# When the best pandemic models are the simplest

**DOI:** 10.1101/2020.06.23.20132522

**Authors:** Sana Jahedi, James A. Yorke

**Author notes:** Corresponding author’s.

## Abstract

As a pandemic of coronavirus spreads across the globe, people debate policies to mitigate its severity. Many complex, highly detailed models have been developed to help policy setters make better decisions. However, the basis of these models is unlikely to be understood by non-experts.

We describe the advantages of simple models for covid-19. We say a model is “**simple”** if its only parameter is the rate of contact between people in the population. This contact rate can vary over time, depending on choices by policy setters. Such models can be understood by a broad audience, and thus can be helpful in explaining the policy decisions to the public. They can be used to evaluate outcomes of different policy strategies. However, simple models have a disadvantage when dealing with inhomogeneous populations.

To augment the power of a simple model to evaluate complicated situations, we add what we call **“satellite”** equations that do not change the original model. For example, with the help of a satellite equation, one could know what his/her chance is of remaining uninfected through the end of epidemic. Satellite equations can model the effect of the epidemic on high-risk individuals, or death rates, or on nursing homes, and other isolated populations.

To compare simple models with complex models, we introduce our **“slightly complex” Model J**. We find the conclusions of simple and complex models can be quite similar. But, for each added complexity, a modeler may have to choose additional parameter values describing who will infect whom under what conditions, choices for which there is often little rationale but that can have a big impact on predictions. Our simulations suggest that the added complexity offers little predictive advantage.

**Author Summary:** There is a large variety of available data about the coronavirus pandemic, but we still lack data about some important factors. Who is likely to infect whom and under what conditions and how long after becoming infected? These factors are the essence of transmission dynamics. Two groups using identical complex models can be expected to make different predictions simply because they make different choices for such transmission parameters in the model. The audience has no way to choose between their predictions. We explain how simple models can be used to answer complex questions by adding what we call satellite equations, addressing questions involving age groups, death rates, and likelihood of transmission to nursing homes and to uninfected, isolated populations. Simple models are ideal for seeing what kinds of interventions are needed to achieve goals of policy setters.

## 1 Introduction

People can sometimes overlook the power of exponential growth, as did the king in an old parable about a servant who had provided a great service. As a reward, the servant asked the king for 64 days of rice, one grain on the first day, two on the second day, four on the third, doubling each day. Initially, the king was delighted with the request, but eventually the king began to understand. The total would be more than 10^15^ kg. So he cut the servant’s head off. Many people would like to cut the head off a pandemic as it slowly spreads, increasing exponentially.

The spread of a virus is reminiscent of Edward Lorenz’s observation that the flap of a butterfly’s wings in Brazil might set off a tornado in Texas. Perhaps a bat in China played the butterfly’s role in the covid-19 outbreak.

A key concept for the transmission of a human infectious disease is “a contact”. A **contact** is defined as an interaction between two people where if the first one is infectious and the other is **Susceptible**, the susceptible person becomes **Exposed** and after a latent period become **Infectious** and finally **Removed**, either through death or recovery with at least temporary immunity. Mathematical models which are used to describe individuals transitioning between the stages susceptible to exposed to infectious to removed are usually called SEIR models or SIR models. For basic references on epidemic modeling refer to [1, 2, 3, 4].

We refer to an **infection’s generation time**, *i.e*., the mean time between being exposed and exposing other individuals, as one **period** [5]. It seems to be approximately 1 week for covid-19, but reliable data is lacking.

For each time period *n*, the average number of contacts that an infectious person has while infectious is called the **contact rate, *β***_***n***_. The contact rate varies with time, depending on interventions like social distancing as well as seasonal fluctuations. Small changes in *β*_*n*_ can result in large changes in the number of cases, due to multiplicative effects [6]. In our models, we assume that infected people are immune after recovery and remain immune for the duration of the simulations.

### Model E (E stands for exponential)

When almost everyone is susceptible, we can model the early stages of an outbreak by as follows,

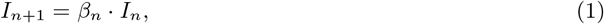

where *I*_*n*_ is the fraction of the population that is infectious in period *n* and *β*_*n*_ is the disease multiplier for each period. When *β*_*n*_ is a constant, we write it as *β*. When *β* > 1, Model E has pure exponential growth, and exponential decay when *β* < 1.

For Model E, *β*_*n*_ is the only parameter that must be chosen by the modeler. We say a model as **simple** if the model’s equation has only one parameter, namely the contact rate. Model E above and Model E+ below are “simple”.

We can think of *I*_*n*_ as being the fraction of the population that is infectious in the *n*^*th*^ generation of the disease. That view is an oversimplification since infectiousness can last a few days or more than a month. This simplification is eliminated in our Model J below. The shortcoming does create small errors in *In*+1 but the errors are small compared with expected errors or uncertainties in the values of *β*_*n*_.

The graphical depiction of case loads can also affect one’s interpretation of the data. The red curves in Fig. 1 show the exponential growth of case rates in Model E as a function of time, assuming that the contact rate remains constant at *β* = 2 (i.e., no mitigation strategy is attempted). The blue curves show the case rates assuming a policy intervention from week 6 to week 11 that temporarily reduces the contact rate. The same data are plotted in two different ways: a linear scale on the left and a logarithmic scale on the right. The plot in the left panel may suggest that the intervention has been a great success, due to the apparent large reduction in infection rates. However, the logarithmic plot shows that the course of the outbreak simply has been delayed by 5 weeks.

**Figure 1:**
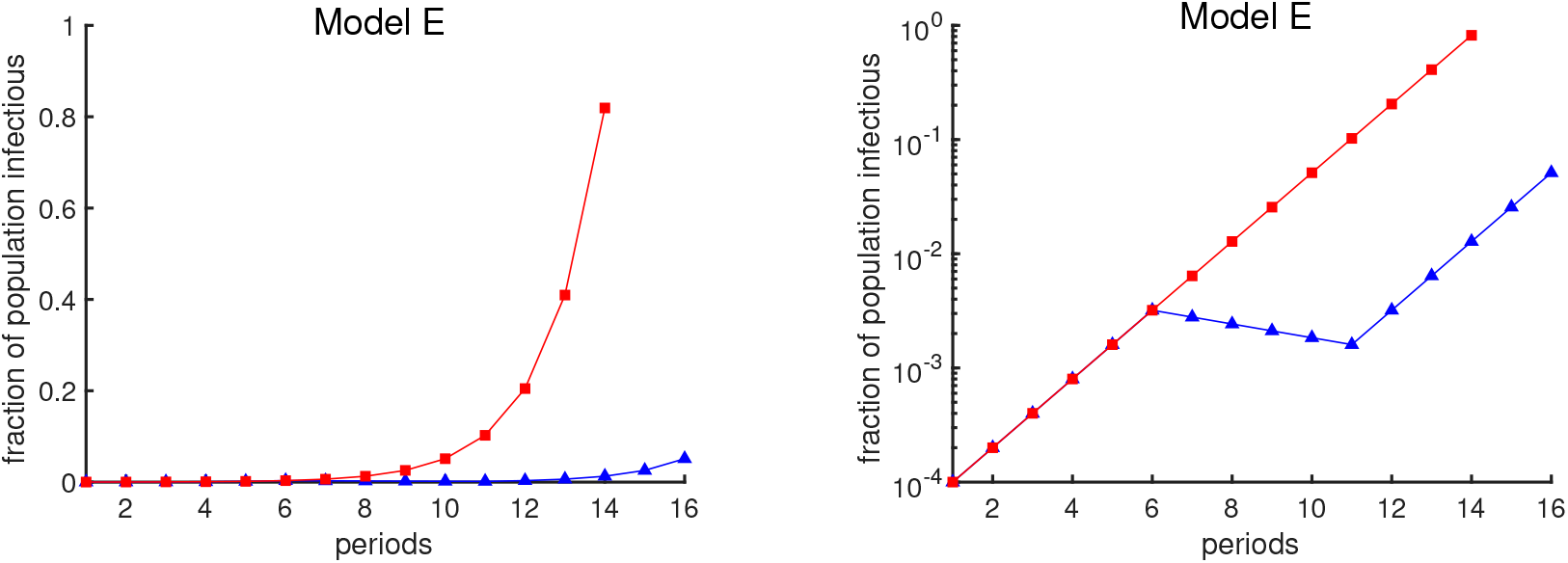
Fraction of the population that is infectious using Model E. Both curves show the fraction of the population that is infectious. One period is average time interval between successive infection generations. The red curve assumes a contact rate of *β* = 2. The blue curve illustrates the effect of an intervention, undertaken between periods 6 and 11, that temporarily reduces the contact rate below 1. The same data are plotted in both panels; the vertical axis is a linear scale on the left and a logarithmic scale on the right.

Infectious diseases can eventually deplete the susceptible population and so do not increase exponentially forever. Hence, we should consider a more realistic model. When proposing a model epidemiologists should have in mind that policymakers should understand a model so that they can make reasonable plans. There are wildly varying sources of advice available. Policymakers are less likely to adopt advice based on models they do not understand and have not participated in [7].

In the following we have proposed two models, a simple model, Model E+, and a slightly complex model, Model J. We will compare the output of these two models and we will show that Model E+ can follow the outbreak as close as model Model J. But any complex model will have many parameters for intra-group contact rates for which there is negligible data, so if plausible choices are used, a wide variety of predictions is inevitable. For example refer to Figs. 5 and 6. Now we introduce the Model E+.

**Figure 2:**
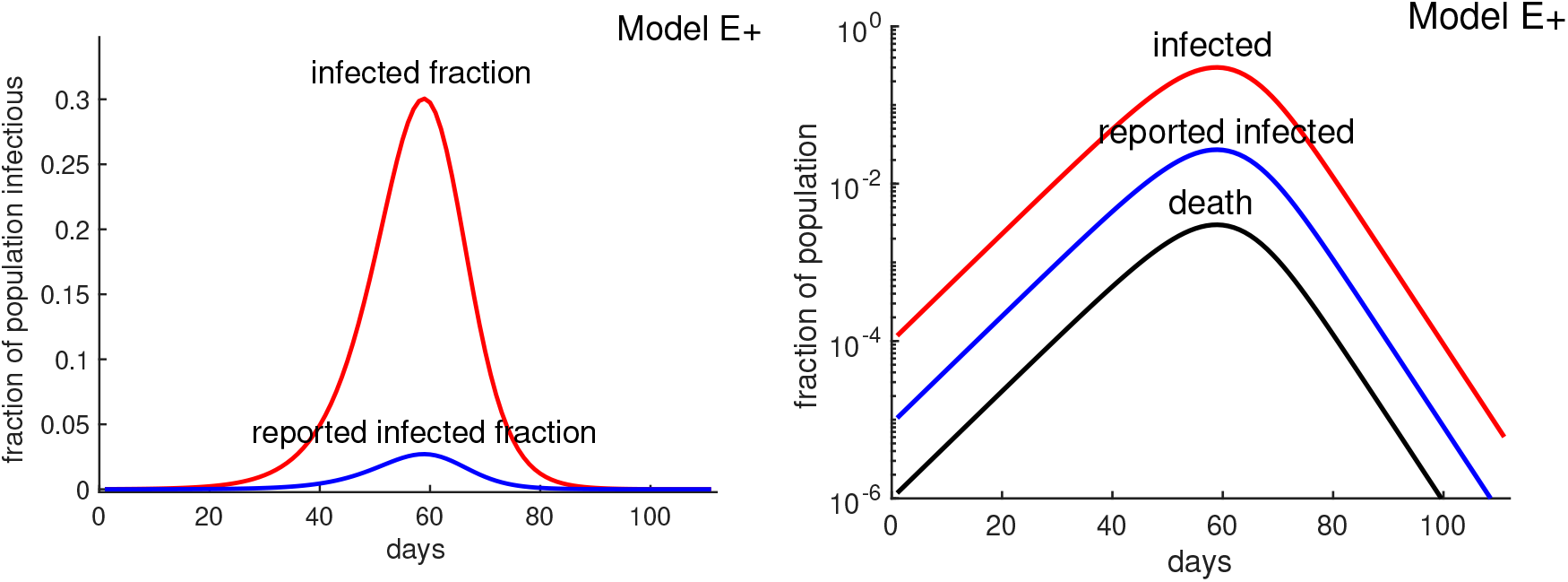
A1l cases vs. reported (confirmed) cases. The higher curve (red curve) represents all infectious cases as a fraction of the population for a hypothetical covid-19 outbreak with *β* = 3 using Model E+. The lower curve (blue) is “reported cases” assuming 9% of cases are reported. The horizontal axis starts at week 0 when the fraction of all cases reach about 10^−4^. The lowest curve (black curve) represents all deaths, assuming death rate is 1%. There is a time delay between being infected and being tested, having the test developed, and then posting the results. Similarly there is typically a delay between being reported and dying. These delays would shift the blue and black curves to the right. We have not shifted the curves and leave it to the reader to decide how much they should be shifted.

**Figure 3:**
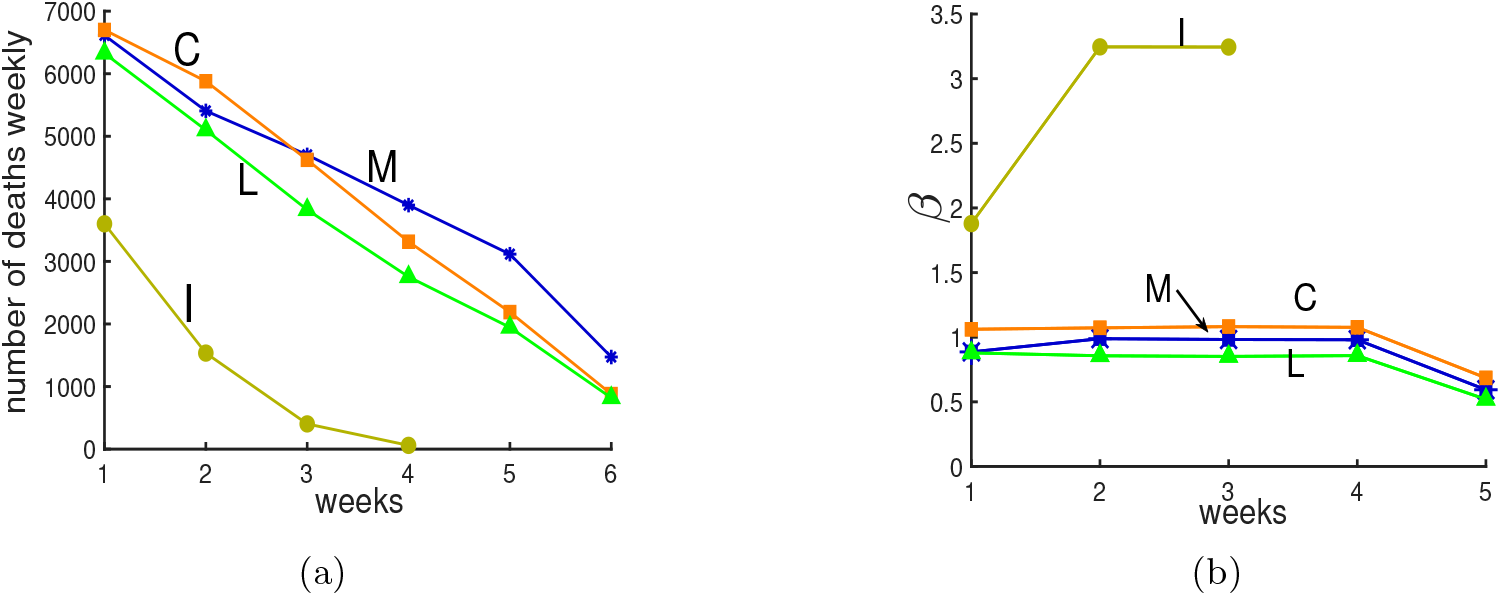
Four simulations from The New York Times. **Panel (a):** Following the beginning of the lockdown in New York City, The New York Times [17] reported on four simulations of deaths with projections for 4 to 6 weeks, shown here with weekly totals. The simulations included large uncertainty intervals that are not shown here. Write *D*_*n*_ for deaths in week *n*. **Panel (b): Contact rates** *β*_*n*_. We assume that deaths are a constant fraction of actual cases, so *D*_*n*+1_/*D*_*n*_ = *I*_*n*+1_/*I*_*n*_. Hence, if one has the initial fraction, *I*_1_, of susceptibles who are infectious and the contact rates *β*_*n*_, then Model E+ will exactly reproduce the curves in panel (a). See the text for a description of the computation of *β* and the values for *I*_1_s.

**Figure 4:**
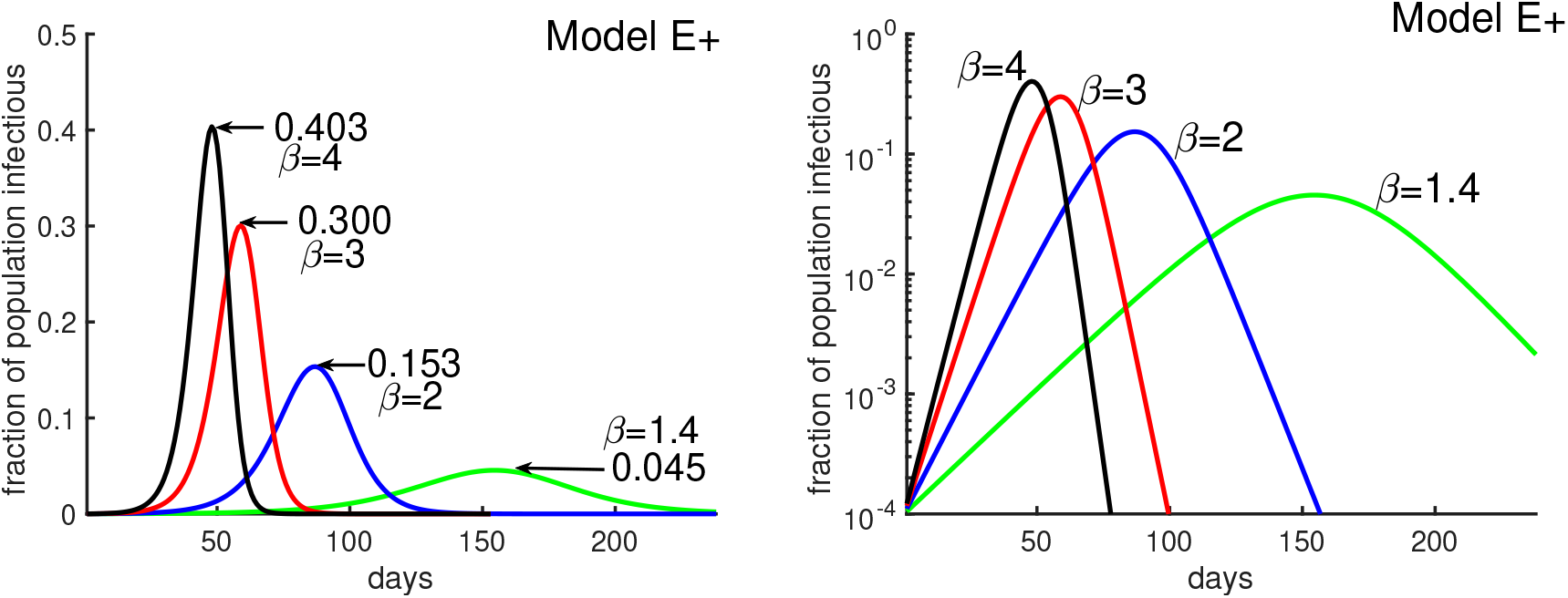
Obtaining daily reports from Model E+. Model E+ is run for ***β* = 2, 3, 4**. Each point on the curves is the infectious fraction for the preceding 7 days, with the horizontal axis being the number of days. The numbers in the graph show each outbreak’s peak-week infectious fraction. To get a smooth curve here and in the previous figure with daily results, we have run Model E+ seven times for each curve, as follows. Early in the outbreak, the fraction infectious increases by the factor ***β*** per week, *i.e*., ***β***^**1/7**^ per day. For each ***β***, the simulation is run seven times with initial infectious ***β***^***j*/7**^, ***j* = 0, 1**, …, **6**, and the plot is shifted to the right by ***j*** days, 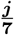 weeks.

**Figure 5:**
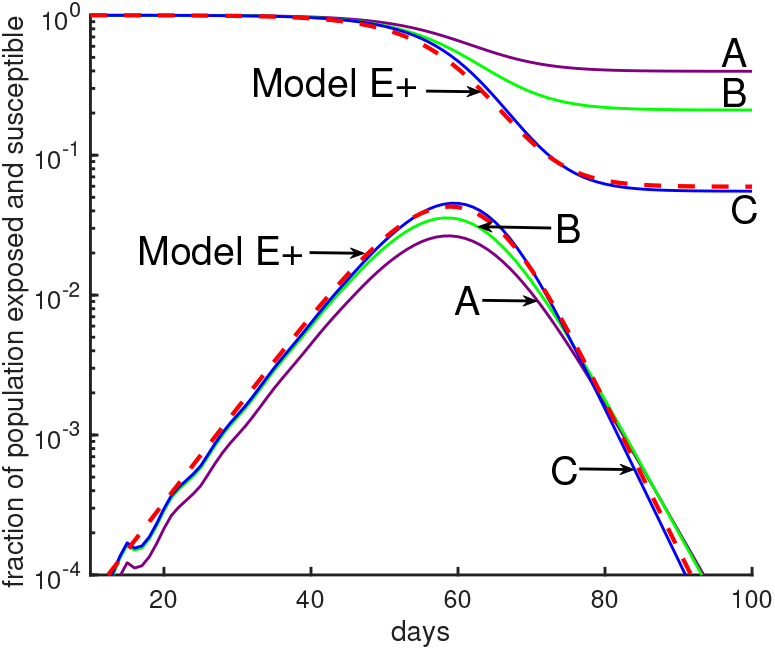
Comparing Model E+’s outbreaks with Model J’s. All simulations in this figure and the next are chosen so that the initial growth rate is a factor of 3 per week. See Table 1 for parameter choices. The initial conditions are chosen so that the 4 curves are initially close together. Changes in initial data will shift a curve to the left or right. **Model E+:** The red dashed curve is for Model E+ with a constant contact rate *β* = 3. **Model J:** The curves *A, B, C*, are from Model J.

**Figure 6:**
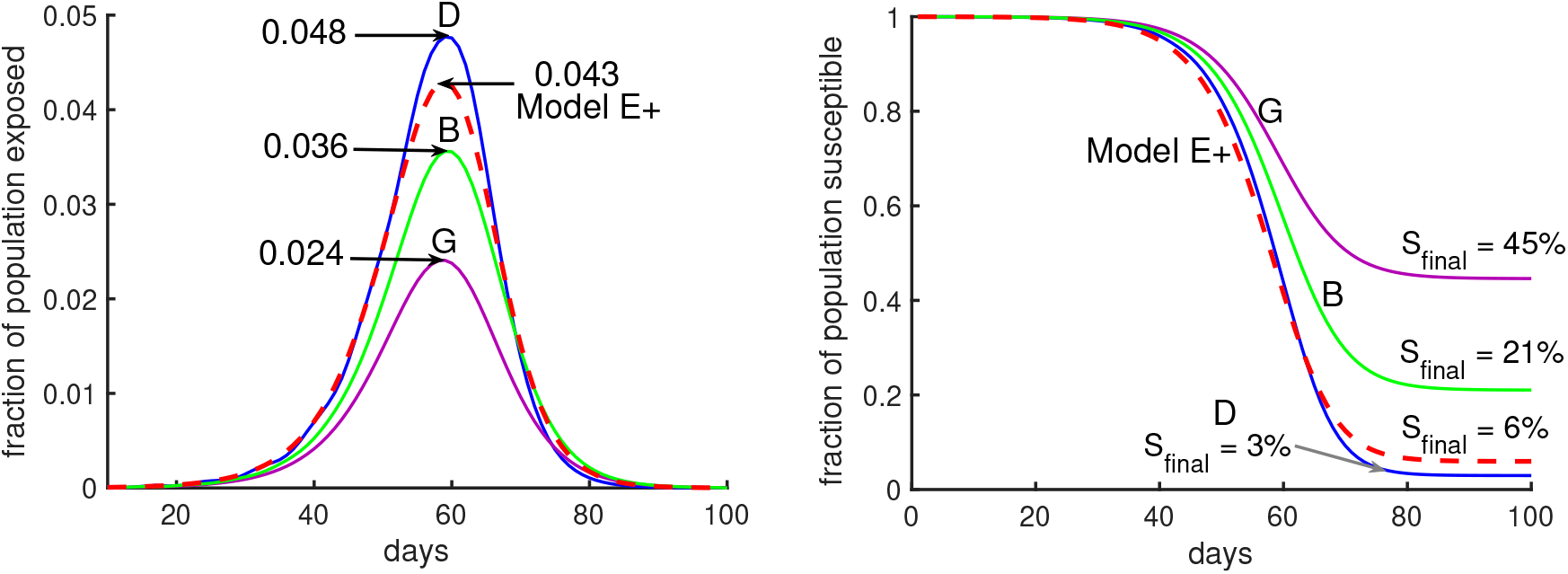
Moderate changes in Model J parameter values make a huge difference. Model J simulations are shown for three sets of parameters, B (green), D (blue), and G (purple). (See Table 1). In both panels the red dashed curve is for Model E+ with a constant contact rate *β* = 3.

### It’s all about contact rates

Model E+ below is designed for a single, “homogeneous” region; *i.e*., it assumes that the fraction of people who are infectious is uniform throughout a region. Specifically, it assumes that at each moment in time the fraction of the population that is infectious is uniform throughout the region. This hypothesis is an approximation since for example, a susceptible who is living with an infectious person has an elevated probability of becoming infected. But we do not know what that probability is, so we neither include that nor any other non-homogeneities in our model.

**Table 1.**

| Figure | Label | color | $\phi_m$ | infectious days | $S_{\text{final}}$ |
| --- | --- | --- | --- | --- | --- |
| Fig. 5 | A | Purple | $m^2$ | | 40% |
| | B | Green | $m$ | 5-9 | 21% |
|  | C | Blue | constant |  | 5.5% |
| Fig. 6 | G | Purple | $m^2$ | 4-8 | 45% |
| | B | Green | $m$ | 5-9 | 21% |
|  | D | Blue | constant | 6-10 | 3% |

We assume the fraction of the population who are infectious at the beginning of the simulation (*n* = 0) should be large enough that immigration of new infectious people is no longer a significant factor in transmission.

While Model E is appropriate when almost everyone is susceptible, the following is more general because it includes the fraction susceptible in week *n*. It is accurate as long as *β*_*n*_ · *I*_*n*_ is small. That means, it is quite unlikely for one person to be contacted by two different infectious people in the same period.

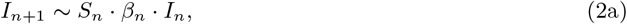

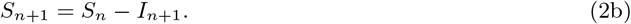

where *S*_*n*_ and *I*_*n*_ are the fractions of the population that are susceptible and infectious respectively in week *n* and *β*_*n*_ is the contact rate in week *n*.

**“Model E+”, a simple SIR model** is our primary model for evaluating intervention strategies. It provides reasonable simulations of an outbreak through the peak as susceptibles become depleted. It corrects the defect of Eq. 2a when *β*_*n*_ · *I*_*n*_ is large, say larger than 0.3. Then the number of contacts is 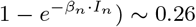. The change to 0.3 might be considered acceptable especially if the true value of *β* is uncertain by perhaps 20%.

### The probability of having no contacts in a particular week

If the expected or average number of events in a time period is *λ*, and the events are independent, the Poisson probability that no events occur is *e*^−*λ*^. The events in question here are contacts in one-week periods. In week *n*, susceptibles average *λ*_*n*_ = *β*_*n*_·*I*_*n*_ infectious contacts, so according to Poisson distribution, the probability of remaining uninfected is 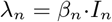, so the probability of becoming infectious is 1−exp(−*β*_*n*_·*I*_*n*_). Hence *I*_*n*+1_ = *S*_*n*_· 1−exp(−*β*_*n*_·*I*_*n*_). The fraction of people in week *n* having a contact with an infectious person is 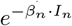 and when that is small (*i.e*., most of the time),

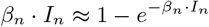

We use the following **“Model E+”**

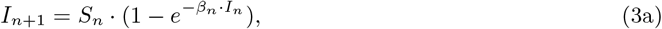

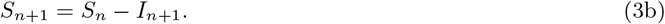

where *S*_*n*_ and *I*_*n*_ are the fractions of the population that are susceptible and infectious respectively in week *n* and *β*_*n*_ is the contact rate in week *n*. People exposed in period *n* are infectious in period *n* + 1 and are removed after that, either becoming immune for the duration of the simulation or dying. People infectious in week *n* + 1 were exposed in week *n* so Model E+ can be considered an SEIR model as well.

Knowing the number of infected individuals, what is the simplest way to predict the number of deaths due to infection? The number of deaths that will result from an epidemic depends on two quantities: The total fraction of the population who will eventually become infected and the fraction of infected people who succumb to disease (the death rate).

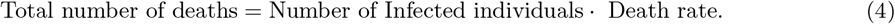

This formula emphasizes how every model’s long-term predictions of deaths will be determined. The covid-19 death rate is likely to be in the range 0.1% to 10%. For a hypothetical population of 100 million, with half getting infected, the model says there will be between 50,000 and 5 million deaths.

While deaths might be estimated during an epidemic, estimating the number of cases is impossible without antibody tests. A detailed model might predict the peak rate of cases, but it cannot use human interaction data to estimate how many of the cases are asymptomatic. Determining the death rate requires both.

### The example of New York City

New York City is heavily dependent on public transportation and perhaps as a result was struck with a massive outbreak. As of May 2020, there were over 185,000 confirmed COVID-19 cases in New York City and an estimated 20,200 deaths. Hence 11% of confirmed cases were fatal. However, it was not known what fraction of total cases were confirmed. If the fraction were close to 1, and if half of New York City’s population of 8.4 million eventually were infected, then according to Eq. 4 the total deaths from the epidemic would be 462,000.

Assuming that the antibody survey results are accurate, the confirmed cases represent less than 9% of the actual cases, which would imply that the death rate is about 1%. Fig. 2 shows that reported infected fraction can be quite different from the actual infected fraction. Thus, if half the population were eventually infected, Eq. 4 would predict about 40,000 deaths. Such estimates are not possible without antibody tests, and it is not possible to know how high the New York City death toll might have risen if “lockdown” policies had not been implemented. Lockdown policies include social distancing, stay-at-home, closing non-essential businesses, and other policies summarized in Sec. 2.

### Model E+, fixing deficiencies of predictions from complex models

As we show below, when you receive predictions of covid cases or deaths from a model, you would really also want to know what fraction of the susceptibles are being infected each time period. The numbers might vary drastically between models, perhaps leading to question a model’s validity.

Following the beginning of the lockdown, The New York Times [17] asked several modeling groups for predictions of covid deaths in New York City. Four groups provided predictions for four or more weeks; they are labeled “C”,”I”,”M”, and “L”, for Columbia University’s team [9], IHME Institution’s response to covid [8], MIT University’s team [11], and Los Alamos National lab’s projections [10], in Fig. 3a. Predictions “C”,”M”, and “L” show progressive decreases in deaths. Prediction “I” shows a more dramatic decrease. Here we would like to determine what makes it different.

### We compute *β*_*n*_ for each prediction as follows

Model E+ concerns infectious fractions, not deaths. Deaths are often used as a more accurate record of how total cases vary than verified cases. For each prediction, we treat the deaths as proportional to the total infectious fraction; *i.e*., deaths_*n*_/*I*_*n*_ = constant or week *n* = 1, 2, …. Hence once *I*_1_ is chosen, all *I*_*n*_ are known. We choose *S*_0_ = 1 and *S*_1_ = 1 − *I*_1_, so *S*_*n*_can be computed from Eq. 3b.

We don’t know *I*_1_, but for each choice of *I*_1_ < 1, we can solve Eq. 3a for each *β*_*n*_ for that prediction. That means that with the computed *β*s, the prediction curves will be reproduced exactly by Model E+. We do not expect *β*_*n*_ to change much during a lockdown, so we find the value of *I*_1_ for which the *β*_*n*_ curve is nearly constant during the middle of the run, yielding Fig. 3b. Those *I*_1_ values for “C”,”I”,”M”, and “L” are 0.1196, 0.6168, 0.0551, and 0.0587, respectively.

Why is Prediction “I” so different from the rest? Does its rapid decline in deaths simply mean the contact rate was chosen extremely low? No. Fig. 3b shows that their contact rate is much higher than for the other three. So why was there such a rapid drop in deaths?

### Our findings

The initial susceptible fraction *S*_1_ required for Prediction “I” is so low that 62% of the susceptibles at the beginning of the first week became infected during the first week, and 96% of the susceptibles were infected in the four weeks shown. Deaths dropped because almost everyone had been infected and almost no one was left to become infected. An alternative hypothesis to explain Prediction “I” is that at this time when we expect the contact rate to be constant, when the other groups provide predictions consistent with nearly constant contacts, the modelers of “I” predicted the contact rates, *β*, would change quickly, dropping rapidly to near 0, thereby stopping the outbreak. No outbreak will persist if the modelers assume the contact rate is near 0.

Any policy setter should want to know why either the infection rates were so high, infecting 62% of the remaining susceptibles in one week, or alternatively what made the contact rates drop to near 0.

**Running Model E+ will exactly reproduce the each of the four prediction curves**, that is, when Model E+ is run with the plotted *β*s together with *I*_1_. We use *S*_0_ = 1. If four complex models produce the epidemic with four different growth rates, why not just choose four different versions of Model E+ with different values of *β*?

This procedure is about a method of analysis where Model E+ allows us to analyze predictions from complex models, without knowing details of the model. It is not an evaluation of the merits of the four models.

### Estimating *β* for the current covid-19 outbreak

Covid-19 began spreading perhaps in early November 2019 and 22 weeks later (April 2) reached 1,201,186 confirmed cases worldwide, which in turn is undoubtedly far below the actual number of people infected. Of course this is noisy data with errors in it. These data imply that the number of confirmed cases grew by at least a factor of approximately 1.9 per week, so the contact rate *β* is at least 1.9. During this time China made major efforts to reduce contacts among its population, making *β* smaller than in areas without such efforts.

We take *β* to be 2 or 3 in most of our simulations. In large cities whose transportation is dominated by mass transit, the growth rate may be much higher. Two recent papers [12, 13] estimated early growth rates as a factor of about 10 or more per week.

Figure 4 is obtained by running four different simulations of Model E+ for contact rate *β* = 1.4, 2, 3, and 4. It shows how the outbreak’s peak depends upon the contact rate. The figure illustrates that a higher contact rate causes a more severe outbreak with a higher and earlier peak. Figure 4 also explains how to produce the outbreak daily using model E+.

### Using Model E+ to determine what happens during an outbreak

The Washington Post reported that from May 25 to June 24 2020, (30 days), a group of states in the United States had increasing covid-19 cases. States had relaxed social distancing following the earlier lockdown. New confirmed cases per day rose from 5,000 to 20,000. That means that during that period, cases increased by a factor of about 1.4 per week. That is, *β* ∼ 1.4 assuming most people were still susceptible. There is a combined population of 121 million for these states, Arizona, Arkansas, California, Florida, N. Carolina, Oregon, S.

Carolina, Texas, Utah, together about 37% of the total U.S. population. Figure 4 says that *if people do not take greater precautions*, we can expect 4.5% of the population to be infected in the peak week for that *β*. We assume the number of confirmed cases is about 1/10 of actual cases. Then the peak *daily*, confirmed cases would peak at 78,000 (∼ 0.045 · 121,000,000 · (1/10) · (1/7)), almost quadruple the June 24 number. Of course the peaks in the different regions might not coincide but this calculation can be thought of as an estimate of the sum of the peak weeks of the different states.

### Model E+ with “satellite” equations

We need models that can inform us about all possible states in our population, and assess which ones matter for shaping the pandemic’s trajectory over time. What will happen to those with pre-existing conditions or older populations or those with different contact rates? How will nursing homes and long-term care facilities be affected? How may will have long-term neurological or other physiological damage from contracting covid-19?

Models E or E+ tell us of the environment people find themselves in. Satellite equations allow us to use additional data about individual reactions to covid and can tell us of the expected impact on individuals. By a **satellite equation** we mean an equation such as Eq. 6 that does not affect the primary system, which in this case is Eq. 3a. The primary system does not depend on the new variables being reported. We emphasize ability to add some complexity without compromising the intelligibility of the original system. We give two examples of satellite equations.

### A satellite equation for predicting deaths

It is well recognized that if hospitals become over-loaded with seriously ill covid patients, the probability that a hospitalized patient will die is elevated. We now outline a scenario that would have to be refined using actual data of hospitalized covid cases. Suppose some fraction *h* of infected people will be hospitalized. To be specific, suppose for example the average hospitalized patient enters a hospital one week after exposure and stays in the hospital for three weeks.

Then the number in the hospital in week *n* would be *h* · (*I*_*n*−1_ + *I*_*n*−2_ + *I*_*n*−3_). Then deaths in week *n* would be predicted by

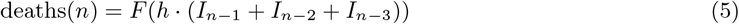

for some nonlinear function *F*. To use this approach, the modeler must determine how long after exposure deaths occur; what *h* is; and what *F* is. Then this satellite equation can be used to compute deaths per week, even though Model E+ does not include deaths.

### A satellite equation for people with a different contact rate

Suppose person A is isolated most of the time, but about once a week, goes out to lunch with a person B, a different B every time. Suppose the luncheon is in a confined space such that if B is infectious, A has a 50% chance of being exposed. What are A’s chances of becoming infected during the outbreak? We proceed by first making some general calculations.

Model E+ uses an average contact rate. To investigate an individual whose contact rate differs from the average person’s, we can adapt Model E+ by adding a satellite equation. A person may have a very low contact rate, due to wearing protective gear or staying at home, or perhaps the rate is very high due to being in crowds frequently. We denote this person’s contact rate in week *n* by *γ*_*n*_, which can vary weekly. Let *P*_*n*_ denote the person’s probability of not being infected by week *n*. We can set *P*_0_ = 1. Here we use Model E+ to determine *I*_*n*_ and *S*_*n*_. We then use *I*_*n*_ in a satellite equation for determining *P*_*n*_,

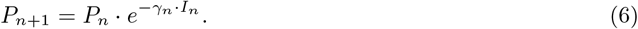

Notice that *P*_*n*+1_ can also be written as follows

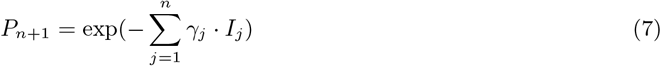

If a person maintains a constant *γ*, then ∑*γ* · *I*_*j*_ = *γ* · (*S*_0_ − *S*_*n*+1_). Choosing *S*_0_ = 1 yields the probability of remaining uninfected at the end of the epidemic,

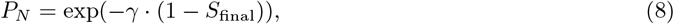

where *S*_final_ denotes the final fraction of susceptible people at the end of the period simulated. To decrease the risk of getting infected, Eq. 8 suggests keeping *γ* low.

Now we return to the life of A. We have in effect said that A has 0.5 contacts per week and is otherwise isolated. That means *γ* = 0.5.

It is likely that some of the B’s that A has contact with will become infected at some point during the outbreak. But A will be exposed only if some B is infectious during the week they have lunch. Suppose at the end of the epidemic 2/3 (= 1 − *S*_*N*_) of the population has become infected. Then Eq. 8 says *P*_*N*_, the probability of remaining uninfected at the end of the outbreak, is exp(−0.5 · (2/3)) = exp(−1/3) ≈ 0.72, or a 28% chance of becoming infected as a result of these lunches.

### A satellite equation for a population with no infections, e.g., a nursing home or an island

Let A denote a relatively isolated population with no cases currently, such as the resident population of a nursing home or a small country or region. Suppose a population A has *N* visitors per infectious period (which we can think of as one week), and the visitors come from a place B that is experiencing an outbreak. Assume the infectious fraction in B is *I*_*n*_ in period *n*. There are different types of visits. Consider the “long-term” visitor, who might be staying more than one period or equivalently is a resident who returned home after a long visit elsewhere. Here “long” means more than one period. A “short-term” visitor might stay for a day.

We can show under reasonable hypotheses that such a person will on average have had half his or her contacts before arrival and half after. The probability that such a visitor is infectious is *I*_*n*_. The expected number of transmissions per period is (*β*/2) · *I*_*n*_ · *N*. Over the duration of the outbreak, the expected number of primary introduced transmissions *T* is the sum as in Eq. 8.

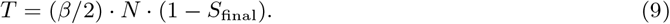

Suppose for example that during the outbreak, 2% (= 1−*S*_final_) of the external population was infected and the number of visitors was 200 per period, and the transmission rate is *β* = 3, then *T* = 6 = (3/2)· 0.02 · 400 primary infections from visitors. If the outbreak has infections transmitted primarily by superspreaders, while most infectious people infect almost no one, then we would expect few new outbreaks but the clusters would be big.

## 2 Factors decreasing the contact rates and their cost

The purpose of a lockdown policy is to cause each person who is infected to infect fewer people. We discuss the United States here but the results are applicable to many countries and to smaller regions.

Before the spring 2020 lockdown period, each infected person infected on average more than 2 people. We discuss policies to control diseases [15] by means other than using drugs.

If we can trust the virus testing statistics, that suggests that most of the potential transmissions were interrupted. The numbers have not dropped as much as the government would like. It would be preferable that there were no new transmissions.

### Costs per case

During the 12-week “lockdown period”, from March through May 2020, we estimate that there were fewer than 10 million infected people. That is based on the following. The U.S. has had less than 10^5^ deaths. Using a 1% death rate, we extrapolate that the number of infected people in the U.S. was less than 10 million, including those whose infections were mild or non-symptomatic.

The costs of the intervention to the US government is at least $2 · 10^12^, the price of the 2020 Stimulus bill called the “CARES act”. We estimate that when we add lost wages and the cost to the private sector, the cost may be about $6 · 10^12^. The cost of interrupting transmissions per infected person is greater than $6 · 10^12^ /10^7^ = $600,000. If the lockdown reduced transmissions per infected person from roughly 3 to 1, then 2 of transmissions were prevented per infected person, at an average cost of $300,000 per prevented transmission. Having such an estimate allows us to ask how many infectious contacts can be eliminated using other strategies.

Eikenberry et al. in [18] discuss the impact of wearing face masks in preventing contacts. Suppose moderately effective masks were available that when worn would prevent 60% of transmissions. If everyone wore them 80% of the time, then 60% of 80% of the cases would be prevented. That is, 48% of all the cases would be prevented. This is not an economics paper so we will not estimate costs of intervention methods.

Many of the interventions below can be thought of as mutually independent, as a first approximation. Each interrupts some fraction of contacts. If an intervention reduces the remaining infectious contacts by some factor *F*_*j*_, then *β*_*n*_ is reduced to *β*_*n*_ · (1 − *F*_*j*_). If *N* independent interventions are applied, then *β*_*n*_ is reduced to

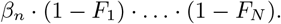

We believe the implementation of each would reduce contacts by some factor 1 − *F*_*j*_. The fraction *F*_*j*_ can only be estimated, would vary over time, and would depend on the implementation.

There are also additive policies. Banning daycare and closing schools both can reduce contacts, but they ban different contacts, so we might say that together the fraction of contacts blocked could be written *F*_*daycare*_ + *F*_*schools*_. Similarly banning sports gathering and religious gatherings would likely be additive.

Each of the following actions or policies would disrupt some fraction *F*_*j*_ of the contacts between infectious and susceptible people.

(F_SD1_) Isolate Infected individuals - those who are not seriously sick to be hospitalized rather than sending them home where they might infect house-mates.

(F_SD2_) Quarantine exposed individuals.

(F_SD3_) Ban large gatherings of people.

(F_SD4_) Recommend social-distancing policies such as the 6 feet / 2 meters rule.

(F_SD5_) Limit travel - only for essential purposes - food, medical, work.

(F_C1_) Close of Non-Essential Businesses (Employees may work from home).

(F_C2_) Close School buildings, educate remotely. Close Day Care facilities.

(F_TMP_) Test front-line medical personnel.

(F_S_) Sterilize frequently touched public surfaces, like shopping cart handles and door knobs.

Wearing masks, gloves and washing hands have long played a large role in preventing the spread of infectious disease. Having all people wear high-quality masks outside the home might be as effective as closing all nonessential business, and would be much less destructive of society and the economy. The magnitudes of the fractions below depend on the protective power of the masks. That power is higher with the highly effective (N95) Masks. Their effectiveness depends in part on their availability, and on educating the population on how to be fitted for the masks.

It is truly strange but true that F_C1_ above could be implemented without requiring essential business employees to wear masks when interacting with the public. A variety of reports spoke of businesses that are actively stopping their employees from wearing masks, perhaps thinking that such masks would intimidate customers.

(F_M1_) Employees wear masks in places of business when they are not alone.

(F_M2_) Expect or require everyone to wear masks in public, indoor (and outdoor?) settings (in public transportation, stores, businesses, clinics). This requirement is more effective when there is an emphasis on making high-quality masks available to the public.

Some factors may be important in saving lives while not falling into the above calculus.

- Supporting isolation of the most at-risk individuals.
- Testing for the virus must be fast and widely available in order for contact tracing to be effective.
- Contact tracing.

When we think of the goal of interventions to be the disruption of contacts of infectious people, we realize that even limited contact tracing can play a role, such as contacting the people who were most likely to be exposed. We should ask what the cost of interception and quarantining exposed individuals is.

## 3 A mildly complex “Model J”

For comparison of conclusions of Model E+ with more complex models, we create our Model J by adding two reasonable and common complications: contact rates that vary from group to group within the population and infectiousness that depends on how much earlier the infected person was exposed. They improve the realism – provided we have data so that we can accurately set the parameters of the model. The downside of this added complexity is that the system is harder to work with and the results are more difficult to communicate to the people who are being advised. To overcome this downside, here we propose an approach that can make complex models more manageable: only one constant must be determined precisely after other constants. Basically, we are more likely to know that A has about twice as many contacts as B than what those actual contact rates are.

The elements of this model are the following.

- Some people have more contacts than others. The population is partitioned into *K* equal-sized groups that can have different contact rates. Usually we take *K* to be 10. *S*^*m*^(*d*) and *E*^*m*^(*d*) are the fractions of the people in group *m* who are susceptible and possibly exposed on day *d*, respectively. Hence, 0 ≤ *S*^*m*^(*d*) ≤ 1 and 0 ≤ *E*^*m*^(*d*) ≤ 1 and initially we can expect *S*^*m*^(1) ∼ 1 for all *m*. Note that we always take *m* to be between 1 and *K*. The number of contacts of people in group *m* per unit of time is proportional to *ϕ*_*m*_. The number of contacts between people in groups *i* and *m* is proportional to *ϕ*_*i*_ · *ϕ*_*m*_. For example we might have *ϕ*_*m*_ = *m* or more generally *ϕ*_*m*_ = *m*^*α*^ for some *α*.
- Infectiousness depends on how long a person has been infected. The “infectiousness” or likelihood of transmitting the infection on day *j* after being exposed is proportional to the constants ***χ***_***j***_. The collective infectiousness of people in group *i* who were exposed *j* days earlier is proportional to *χ*_*j*_ · *E*^*i*^(*d* − *j*). Hence, the collective infectiousness also depends on the contact rates.
- *E*^*m*^ (*d*) is the fraction of susceptible people in group *m* being exposed on day *d* by people from group *i* who were infected *j* days earlier. *E*^*m*^(*d*) is proportional to

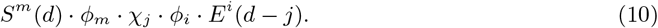

Summing over all *i* and *j, E*^*m*^(*d*) is proportional to

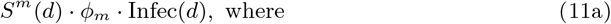

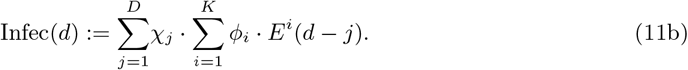

The term Infec(*d*) is proportional to the size of the total infectious population on day *d*; therefore, it is proportional to the level of danger to the community on day *d*.
- The novel feature of this model is that only one parameter must be determined precisely in the model. We can make Eq. 10 exact rather than proportional by multiplying it by the appropriate value, *J*. The actual transmission rate is

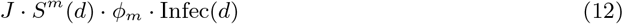

*J* is independent of *m, i*, and *j*. Below we show how to select *J* so that the outbreak has an initial growth rate *β*. Typically we choose *β* = 3. *J* is time independent if there is no intervention, but interventions can make *J* = *J* (*d*) depend on day *d*.

In summary, the resulting **Model J** follows. For each group *m*, 1 ≤ *m* ≤ *K*, we have

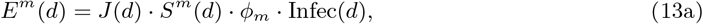

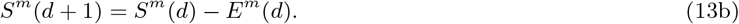

### Tuning Model J to achieve a specific growth rate *β*

David Adam, an editor of *Nature*, reported in a News article [19] that two very different models produced nearly estimates of the number of deaths that differed by 1%. That similarity gives the illusion of reliability and robustness, but, in fact, the fraction of infected people who die is unknown. As shown by the evolving New York City data, estimates of the total number of deaths can be uncertain by a factor of 10. Modelers must tune their models to reflect reality. In simple models, that means choosing *β*_*n*_ and the initial fractions of the population that are infected and perhaps susceptible (for Model E+). All these choices should be under discussion with the policy setters.

Suppose we are observing an outbreak that grows by a factor of *β* for a time period *τ*, and suppose we want to tune Model J so that it has the same growth rate initially. There is a value of *J* such that for any constant C, the following is a solution of Eq. 13a:

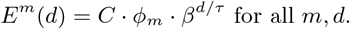

Substitute *E*^*m*^(*d*) into Eq. 13a with *S*^*m*^(*d*) = 1 for all *d*. Then after factoring out *β*^*d*/*τ*^, for each *m* we obtain the following equation for *J*.

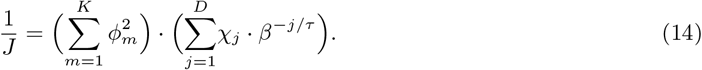

It is independent of *m, j, C*, and *d*.

### Parameter sets for Model J

The main purpose of introducing Model J is to show how many parameters there can be. But it is also to show how a model with relative parameters can be tuned by choosing one parameter, *J* for the whole system.

Five examples illustrate the behavior of the model: A, B, C, D, and G in Figs 5 and 6. For each, we divide the population into 10 groups that can have different contact rates, and exposed people are infectious for 5 days, and they are equally infectious for each of those days. Which days they are infectious depends on the case, see table 1 for more details. For most cases we set the five days to be 5-9 days after exposure.

When there is no intervention, for example in simulations of Model J in Fig. 5, the parameter *J* is chosen constant (independent of *d*) so that initially the epidemic grows by a factor of 3 per week. That corresponds to using *β* = 3 and *τ* = 7 days in Eq. 14.

The outbreaks of Model J can be quite similar to Model E+’s, as seen in Fig. 5, but reasonable changes in the *ϕ*s and *χ*s can produce big variations in the predictions of the Model J as seen in Fig. 6.

Figure 5 shows outbreaks from our Model J (Eq. 13). We draw the reader’s attention to the fact that there is more variation between the three Model J curves than there is between their average and the Model E+ curve. We suggest that it is quite difficult to determine which of the three Model J simulations best represents reality. We have seen in Fig. 4 that the outbreaks depend strongly on *β*. A small change in *β* is more significant than whether Model E+ or Model J is used, and *β* has been difficult to determine for actual populations. Hence we believe little is gained by using more complex models for setting policy. The cost of complexity would be a loss of intelligibility for most policy setters.

One model by researchers from Imperial College is described as having 15,000 lines of code, see the url Code Review of Ferguson’s Model [14]. That model has a great deal of geographic detail, with infections transmitted from region to region. Uncertainties in transmission rates make prediction like trying to predict the chaos of billiards when you don’t know the properties of the billiards.

In Fig. 6 we show two cases where the **mean infectious time** is either day 6 after exposure (using infectious period days 4-8, Case G) or day 8 (using days 6-10, Case D). See Table 1. In each case, Model J has been calibrated though the choice of its parameter *J* so that the initial rate of growth is 3. Initial conditions have been chosen so that during the early parts of the outbreak, the curves are aligned. Small changes in the initial fraction infected shifts each curve to the left or right without otherwise changing it.

Our main choice of *χ*s assumes people are infectious for five days, days 5 through 9, and they are equally infectious each of those days; that is *χ*_*j*_ = 0 except for *j* = 5, …, 9 when *χ*_*j*_ = 1.

Model J is useful because we can test its sensitivity to a variety of choices of parameters. In such a comparison, the exponential growth rate early in the epidemic should be the same for both models.

We find the differences in predictions between Model E+ and Model J are small compared with the uncertainty of the parameters such as the initial growth rate or the death rate.

### Reasonable variations in choices of parameters yield a big impact on Model J outbreaks

In practice we don’t know how much difference there is between the personal contacts of people nor do we know precisely when the typical infected person is infectious.

Suppose we want to model a city. We have to choose values for *ϕ*s and *χ*s. Fig. 6 shows outbreaks using several choices. It would be difficult to know which choice is most appropriate. However in the right panel of Fig. 6, the fraction of the total population remaining uninfected at the end of the outbreak, *S*_final_, is 15 times higher for case *G* (purple curve) than for case *D* (blue curve). Case *D* has a peak that is twice as high as case *G*’s.

## 4 Results

### The effect of Intervention

We model two policies to mitigate the severity of the outbreak using Model E+ and Model J.

1. Policy I : During an intervention period, the contact rate is reduced by a fixed factor.
2. Policy II: We reduce the contact rate weekly to keep the maximum fraction of infectious people below a certain threshold, so as not to exceed the capacity of the healthcare system.

### Policy I: A short-term intervention

We investigate the effect of a 6-week intervention during which the effective contact rate *β* is reduced from 3 to 1.6. Such interventions in an Influenza epidemic can delay the growth of the epidemic [16]. Our simulations show this delay for an early intervention. By an early intervention we mean intervention be implemented more than three weeks before the uncontrolled outbreak would peak. Our simulations suggest that an early intervention only delays the epidemic (blue curves in Figs. 7a and 7b). A late intervention is being implemented one week before the uncontrolled outbreak peaks. A late intervention will not have a very useful impact on reducing the peak, but it will increase *S*_final_ slightly (purple curves in Figs. 7a and 7b). According to our simulations, a policy I type intervention has an optimum impact on reducing the severity of outbreak if it starts either two or three weeks before the uncontrolled outbreak peaks. An intervention which starts two to three weeks before uncontrolled outbreak peaks is called an intermediate intervention. For Model E+ when *β* = 3, the uncontrolled outbreak peaks at week 12, with a peak ∼ 28% and S_final_ ∼ 6%. An intermediate intervention can reduce the peak by 50% and the fraction of people who do not experience the disease will increase by more than 50%. (Golden curves in Fig. 7a and 7b).

**Figure 7:**
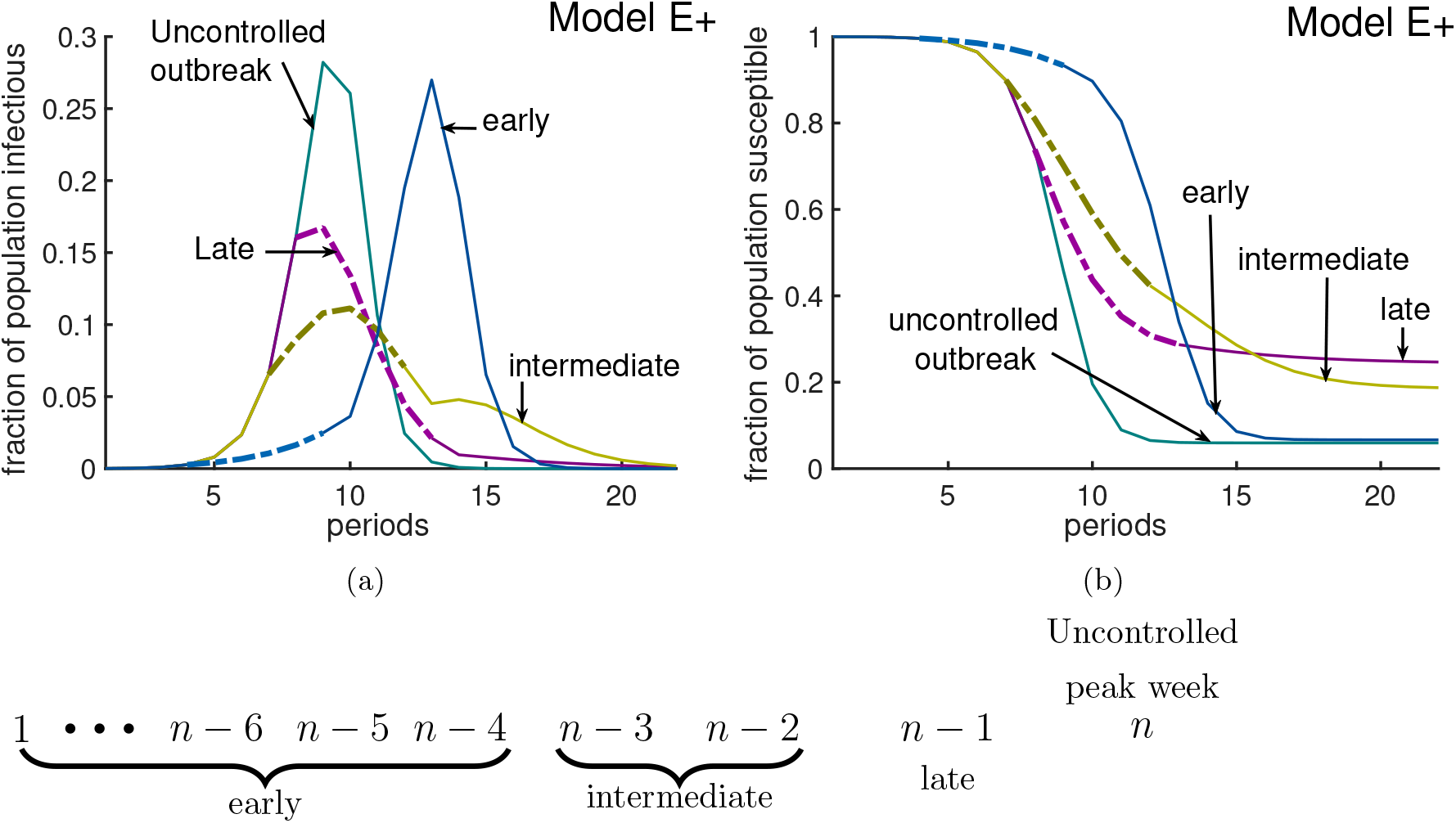
Timing an intervention for optimal effect. In this simulation, the rate of contact is *β* = 3 before and after intervention, during an intervention *β*^∗^ = 1.6. The Green curves represent the uncontrolled outbreak without any intervention, the peak is about 28% and S_final_ is about 6%. Panel (a) represents the fraction of infectious individuals and panel (b) shows the susceptible fraction of population. Here we have implemented 3 different 6-week long interventions. An early intervention (blue curves), starts at week 5 is not very interesting compared to an intermediate intervention. If the uncontrolled outbreak peaks at week *n*, then an intervention will be called early, intermediate, and late if it is implemented at least 4 weeks earlier than, 2 to 3 weeks earlier than and exactly one week before week *n*, respectively. As the figure illustrates an early intervention only delays the outbreak. A late intervention (purple curves), starts at week 8 considerably increases the fraction of people who remain uninfected at the end of outbreak. A middle intervention (golden curves) should be applied two weeks before the uncontrolled outbreak peaks. According to this figure the optimal effect, which is 50% reduction in peak and 50% increase in fraction of left susceptible, is achievable by the middle intervention.

One might think that harsher restrictions will stop an outbreak. The three panels of Figure 8 show a variety scenarios for interventions, all lasting from weeks 7-12, during which the contact rate is reduced. Panels (a) and (b) show Model E+ simulations. The two in (a) are among the seven in (b) but the vertical scale in (b) is logarithmic. Panel (c) is the same as (b) except that Model J is used. In all three panels, the numbers tell the value of *β* used during the interventions.

**Figure 8:**
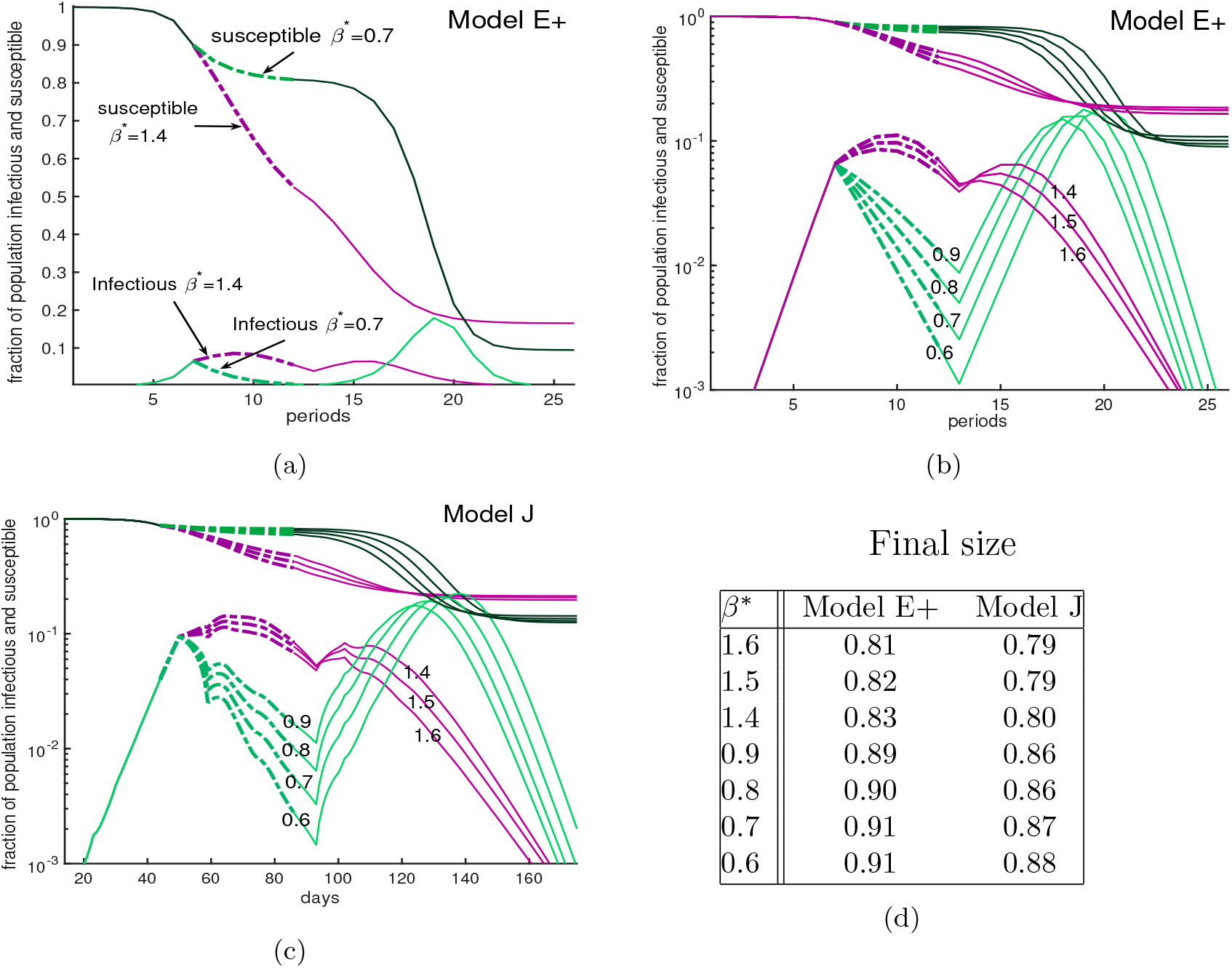
A severe intervention is more likely than a mild intervention to be followed by a large peak. Outside the intervention period, *β* = 3. During intervention the contact rate is shown by *β*^∗^. The curves are dashed during the intervention period. **Panel (a):** This figure compares a mild intervention with a harsh intervention using Model E+. The uncontrolled outbreak with no intervention (not shown) has a peak about 28% and S_final_ ∼ 6%; *i.e*., 94% of the population is infected during the entire outbreak. Mild intervention, *β*^∗^ = 1.6, raises S_final_ to ∼ 25%. The severe intervention raises the S_final_ to ∼ 18%. **Panel (b) and (c)**. We show an outbreak with a 6-week intervention with a variety of contact rates for both Model E+ (panel b) and Model J (panel c). The intervention period begins when the susceptible populations *S*(*d*) is ≈ 0.9. The vertical axis is showing the fraction of the population susceptible (upper curves) in week *n* for Model E+ or in day *d* for Model J. The lower curves are *I*_*n*_ for Model E+ and Infec(*d*) for Model J. Mild interventions (magenta) reduce *β* to 1.4 to 1.6 and severe ones (green) reduce *β* to 0.6 to 0.9. Model J changes *β* by changing *J*_*β*_ using Eq. 14. For Model J, exposed individuals are infectious on days 7 through 12 and they are equally infectious. **panel (d):** The final size are shown for both Model E+ and Model J. Final size is the total fraction infected during the entire outbreak. According to table in (d) the harsh intervention results in more infected cases during the entire outbreak than the mild one.

**Figure 9:**
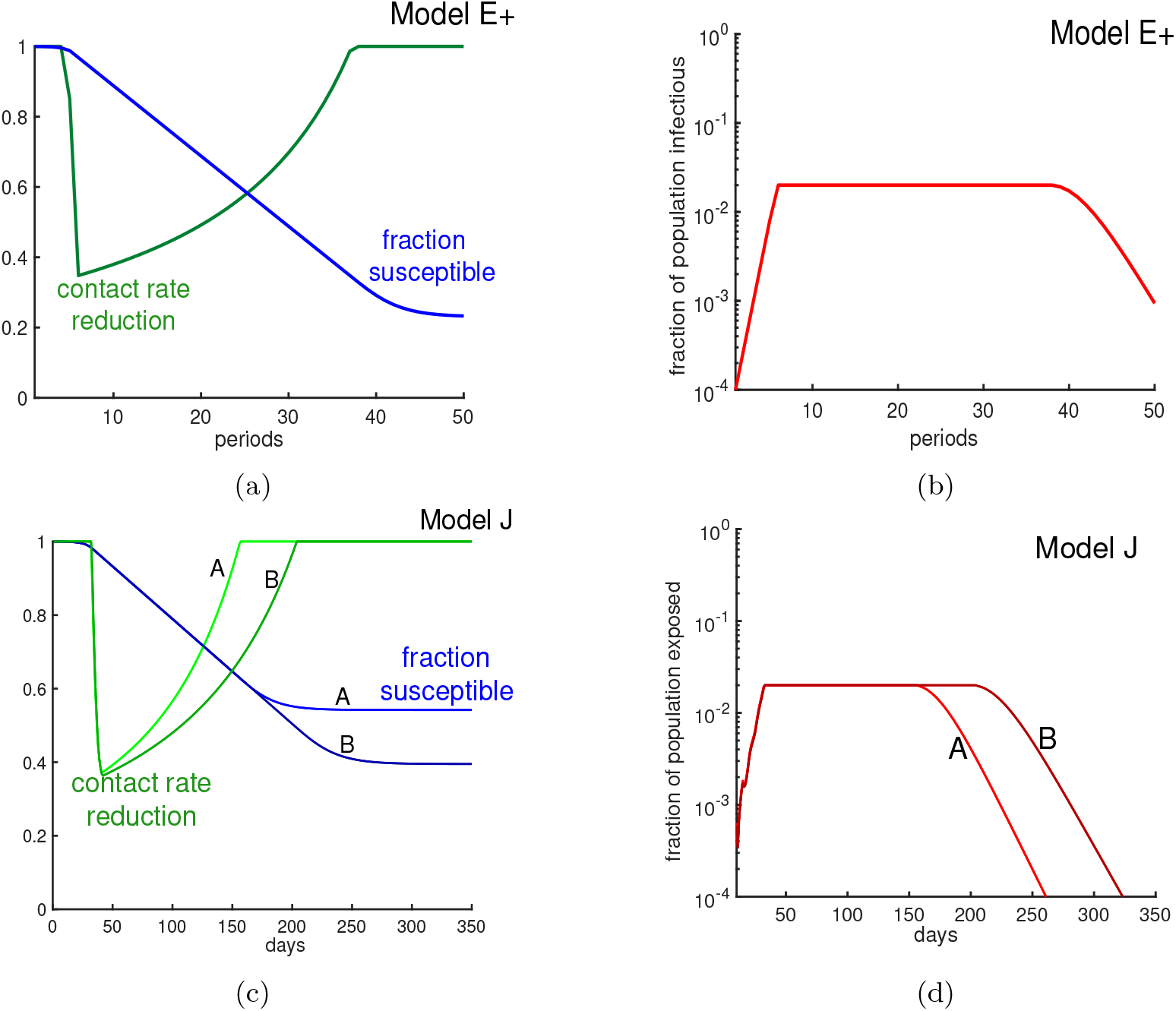
Limit *β* to limit the infectious fraction. Here we change the contact rate weekly in order to keep the fraction of infectious individuals below 2%. The green curve in panel (a) and panel (c) are representing weekly contact rate reduction. As the panels (b) and (d) illustrate, the fraction of infectious individuals never goes beyond 2% per week. Panel (b) is derived from Model E+ and panel (d) is derived from Model J. Alternative strategies include minimizing the contacts of those with risk factors.

The green curves show a severe intervention for various choices of *β* from 0.6 to 0.9 while purple has values 1.4 to 1.6. All reduce the initial peak of infections below what would have occurred if there was no intervention. The milder interventions here allow the number of infections to grow and approach a herd immunity. The severe interventions (green) cause a bigger drop in cases, which is follow by a large peak.

The mild interventions result in more cases during the intervention but fewer total, because the population reaches a herd immunity level during the intervention. The most important point of this figure is that in this complex simulation, Model E+ displays the same behavior as Model J.

### Policy II: Put a cap on the number of infectious cases

When facing a new pandemic, due to our lack of knowledge, even experts cannot predict the longevity of the pandemic without a vaccine.

As an article in New York Times reported [17], most models do not suggest how long a lockdown should continue, or under which conditions we can get back to our regular life routine. A common concern is that a long recession will cause a huge economic downturn. Then what metrics should governors use to allow people to return to their work?

Our policy II allows authorities to keep the maximum fraction of people who are infectious under a certain level, varying interventions week by week to achieve desired contact rates. It could be applied by putting a cap on the fraction of infectious people or exposed people, reducing the pressure on the hospitals.

### Tuning Model E+ to limit the fraction infectious weekly

Let us assume we want to keep the maximum fraction of infectious people at target level. When Model E+ first predicts *I*_*n*+1_ ≥ target, we solve for *β*_*n*_ to get *I*_*n*+1_ = target. Eq. 3a yields 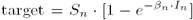. Solving this for the weekly contact rate *β*_*n*_ gives

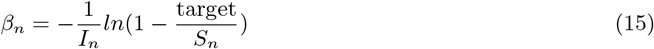

In Fig. (9b), we keep the number of infectious individuals lower than 2%. As Fig. (9a) illustrates by applying this policy about 23% of individuals remain uninfected at the end of outbreak, which has been increased by almost 8% in comparison with the otherwise uncontrolled outbreak.

### Tuning Model J to limit the fraction exposed daily

We will denote the fraction of the total population that is exposed on day *d* by *E*_all_(*d*). Hence,

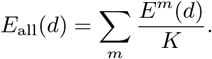

To keep *E*_all_(*d*) below some daily exposure rate that we call “target”, we have to adjust *J* dynamically and in practice that means interventions are needed. When *E*_all_(*d*) would exceed that level without intervention, we decrease *J* (*d*) as follows:

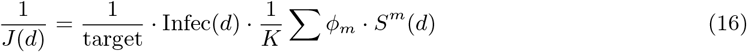

In Fig. (9d) by using Eq. 16 we keep the total exposed under 2% weekly. To make Model E+, which has a time step of 7 days, comparable with Model J, which has a time step of one day, we plot the exposures per week for both models. Note that for Model E+ exposures in week *n* equals the infectious in week *n* + 1. As Fig. (9d) illustrates depending on the set of chosen parameters the desired result can be achievable in shorter (case A) or longer (case B) time. When facing a pandemic, there are lots of uncertainties in the choice of parameters. Choosing the wrong parameters means obtaining a wrong prediction. Reporting all possible predictions from all possible choices results in a very uncertain prediction.

## 5 Discussion

### What policy setters should want

During an outbreak, policy setters will have a variety of advisers giving advice. Models that are understandable have a clear advantage, especially when delivering unpopular advice.

As we show in the introduction’s “Model E+, fixing deficiencies of predictions from complex models”, complex model predictions may fail to provide key determinants of their predictions, like the fraction of susceptibles infected each period.

The policy setters should want to know what assumptions determine the model’s behavior. That includes explanations of how the parameter values are determined or estimated. First the policy setter should understand simple models. Model E is sufficient when the great majority of people are still susceptible.

Simple models for an outbreak in a limited region are applicable to many other actual and potential pandemics.

### Limitations of modeling

Complex models seem to benefit from the common belief in the power of data. In a world-wide outbreak there is plenty of data. But when dealing with a deadly disease, reliable experiments are few. Much of the data is worthless to modelers. Complex models can include many parameters that are precise, such as the typical traffic on highways or numbers of people taking mass transit. Their weaknesses is the uncertainty in the parameters that describe transmissions between groups.

Complex models are likely to have parameters that describe how many contacts people have in the many specific situations. The most accurately known parameter for transmissions between groups of people is the value of *β* (or *R*_0_). The rate *β* is not estimated by examining the individual contacts between people. It is estimated from large amounts of data about the growth in the numbers of infected people or hospitalizations or deaths. This most accurate number is likely quite uncertain. If *β* appears to be 2.5, it may be 2 or 3.5. But there is much less data to determine the transmission parameters or contact rates between subgroups. Hence complex models must be saddled by transmission parameters between subgroups that are less accurate than *β*.

While people are infectious and perhaps asymptomatic, they can encounter many people. How can a model predict how many of these encounters would constitute a contact that transmits infection? We may know the fractions of the populations that are in their 40’s or 60’s but we don’t know the contact rates for disease transmission between the two groups.

The following example illustrates how epidemics are chains of events whose probabilities are hard to compute because each interaction modeled requires its own (unknown) contact rate. The uncertainty in that contact rate exceeds the benefit of including the interaction in the model.

### Consider a chain of contacts

When two people live together, and one, “A”, becomes infected, what is the probability that the other, “B”, will be infected? If an answer to this is obtained in one country, will it be valid for others? New York City apartments might be different from Arizona ranches. Without considerable accurate data, there is a large range of possible parameters values that must be investigated when making predictions with no way to choose between them.

Suppose the probability of the housemate, “B”, being infected is between *P*_1_ and *P*_2_with *P*_1_ < *P*_2_. The “uncertainty factor” can be said to be *P*_2_/*P*_1_.

If “B” now is infectious and *might* ride on a standing-room-only bus or train, how many will become exposed as a result? Suppose the number is between *N*_1_ and *N*_2_ with *N*_1_ < *N*_2_. The uncertainty factor is about *N*_2_/*N*_1_. Hence the possibly infectious person “B” might take a ride on crowded public transportation. (We may have a good estimate as to how often that happens.) The uncertainty factor of how many people are likely to be infected is *P*_2_/*P*_1_· *N*_2_/*N*_1_ by “B” during the ride.

Next suppose that some uncertain fraction *F*_2_/*F*_1_ of the people on the bus go to a football game or similar crowded event. Each infected person is likely to infect an uncertain number *M*_2_/*M*_1_ of attendees. Again the uncertain factors multiply.

The uncertainty factor for this chain of events could be estimated as (*P*_2_/*P*_1_) · (*N*_2_/*N*_1_) · (*F*_2_/*F*_1_) · (*M*_2_/*M*_1_). These ever-compounding uncertainties will contribute to errors in the average contact rate *β*. Since the above chain of infections involves 3 stages of transmissions, it creates uncertainty in *β*^3^ if we are using this chain as part of an estimation of *β*.

Technically, one might argue that we should be discussing standard deviations or variances of each event of the chain. That would require assigning a probability distribution to each step. Then the variance of the log of the number of people infected in such a chain of events is the sum of the variances of logarithms. But we prefer to keep the discussion less technical.

### Tuning complex models so that it gets “reasonable” predictions

Of course all the uncertainties should be tuned so that the resulting *β* is in agreement with the data. Our Model J is designed with that in mind. To minimize the problem of tuning, we use a single parameter *J* which is introduced in Eq. 13. *J* (*d*) could be chosen to be constant to represent an outbreak without interventions. Even then the relative parameters like *χ* and *ϕ* must be selected. The numbers *J* (*d*) must be set by the modeler so that the overall outbreak meets expectations; *J* (*d*) can change with *d*, reflecting what the modeler thinks will happen as a result of interventions. That includes estimates of how people will react to imposed conditions.

Undoubtedly modelers calibrate their complex models against reality as an outbreak progresses. That means they alter the uncertain values of numbers like *P, N, F, M*, … so that a *β* is obtained that agrees with observations. The alterations raise the question of why someone should use a model that requires so many estimates? Why not just pick a model like Model E+ in which the outbreak depends only on one parameter, the contact rate *β*? Different choices lead to different realizations of the course of the outbreak.

### Adding features to complex models

It is attractive to include many features when modeling an outbreak. Each added feature enhances the appearance of reality. There are many possible refinements. Models can split the population into many small groups, perhaps by age, sex, their locations, or population density. Models may also include gatherings for sports, music, movies, religion, politics, or holiday and beach festivities. How many people at such a gathering will be exposed by one infectious person? It depends on the type of gathering, and in any case the answers are not known.

A model must describe how each feature or category affects transmission. Modelers caution against including phenomena with unknown values [20]. Often no data is available. Then the modelers must build in their speculations.

Before a model’s predictions are released, its numbers will be adjusted so that the epidemic’s growth rate will be in agreement with the observed growth rate. The current growth rate is hard to estimate because of the lack of consistent random screening data. However, the current growth rate is the aspect for which there the most data is available, even if that data is inadequate. It cannot be derived from collections of model features for which there is less data.

We believe that the fewer the features a model has, the more intelligible it and its assumptions are. We find that the difference between Model E+ and Model J is small compared with changes in *β*.

### Our approach

To run a simulation using Model E+, Eq. 3a, a modeler chooses *I*_0_ and *β*. Those who set policy can choose what *β* should be, then can choose and vary policies aimed at achieving the target *β*.

We do not predict the future of an outbreak. Instead, we only discuss what will happen for different choices of *β*. We select *β* directly. That will determine when the outbreak peaks, if a large enough fraction of the population is infected. The policy maker makes choices that determine *β*_*n*_. The contact rate will vary week by week as interventions change the contact rate.

## 6 Conclusion

### Estimating *β*_*n*_

We believe the most important factors in predicting the severity of an outbreak are the contact rate, the hospitalization rate, and the death rate. A higher contact rate leads to a higher peak. Modeling interactions between people will not reveal the death rate. The contact rate *β* can be estimated from data. The overall contact rate of the population is the easiest interaction parameter to evaluate.

### Policies I and II

Suppose the goal is to reduce the susceptible population so as to approach herd immunity. We offered two different control policies. In policy I we only apply lockdown restrictions only for a short period of time and then we relax the restrictions. All our simulation is done only for 6-week long interventions. Of course a longer intervention will have a better effect on reducing the unpleasant effect of the outbreak, but also a higher economic cost.

The benefit of policy I might be to provide enough time for supplying the medical establishment. If that is the goal, it would be best to apply inexpensive interventions early, like requiring business personnel to wear masks and applying social distancing. In the United States, during the covid outbreak, nonessential businesses would not have been closed (high economic cost) if the use of masks (low cost) was recommended.

According to our simulations, an excellent control for policy I is a middle intervention which should be implemented two to three weeks before the uncontrolled outbreak peaks. It can reduce the peak by 50% and increase the people who remain uninfected at the end of outbreak by 50%. A harsh intervention will be more likely to be followed by a larger peak than a milder intervention.

When we face a situation in an outbreak where we lack healthcare staff and lack space in our hospitals, it is especially important to keep the total fraction of individuals who are infectious under a target percentage. Then we suggest implementing policy II in which authorities change the social-distancing policies weekly to maintain the maximum number of infectious individuals under the bearable target.

### Interventions, their cost and their effectiveness

When choosing between interventions, it would be advantageous to evaluate each intervention and estimate how many contacts will be disrupted by its implementation and the cost per interrupted contact. The costs include jobs lost, businesses being closed temporarily, or permanently. Interrupting contacts with people at high risk has extra importance. Complex models and simple models with satellite equations are designed to take into account some of these types of contacts. The data for such may not allow us to use the model to determine *β*, but it may be beneficial to categorize interventions by cost/benefit ratios. Many of the features of complex models can be achieved using satellite equations with Models E or E+.

## Data Availability

All the data that has been used here can be available upon request.

## Acknowledgments

We thank Ishan Saha, Eric Kostelich, Aleksey Zimin, Louise Raphael and James Watmough for their input.

